# Advancing the prediction and understanding of placebo responses in chronic back pain using large language models

**DOI:** 10.1101/2025.01.21.25320888

**Authors:** Diogo A.P. Nunes, Dan Furrer, Sara Berger, Guillermo Cecchi, Joana Ferreira-Gomes, Fani Neto, David Martins de Matos, A. Vania Apkarian, Paulo Branco

## Abstract

Placebo analgesia in chronic pain is a widely studied clinical phenomenon, where expectations about the effectiveness of a treatment can result in substantial pain relief when using an inert treatment agent. While placebos offer an opportunity for non-pharmacological treatment in chronic pain, not everyone demonstrates an analgesic response. Prior research has identified biopsychosocial factors that determine the likelihood of an individual to respond to a placebo, yet generalizability and ecological validity in those studies have been limited due to the inability to account for dynamic personal and treatment effects—which are well-known to play a role. Here, we assessed the potential of using fine-tuned large language models (LLMs) to predict placebo responders in chronic low-back pain using contextual features extracted from patient interviews, as they speak about their lifestyle, pain, and treatment history. We re-analyzed data from two clinical trials where individuals performed open-ended interviews and used these to develop a predictive model of placebo response. Our findings demonstrate that semantic features extracted with LLMs accurately predicted placebo responders, achieving a classification accuracy of 74% in unseen data, and validating with 70% accuracy in an independent cohort. Further, LLMs eliminated the need for pre-selecting search terms or to use dictionary approaches, enabling a fully data-driven approach. This LLM method further provided interpretable insights into psychosocial factors underlying placebo responses, highlighting nuanced linguistic patterns linked to responder status, which tap into semantic dimensions such as “anxiety,” “resignation,” and “hope.” These findings expand on prior research by integrating state-of-art NLP techniques to address limitations in interpretability and context sensitivity of standard methods like bag-of-words and dictionary-based approaches. This method highlights the role of language models to link language and psychological states, paving the way for a deeper yet quantitative exploration of biopsychosocial phenomena, and to understand how they relate to treatment outcomes, including placebo.

## Introduction

The placebo effect is a widely studied clinical phenomenon, by which an inert substance can strongly influence psychological and physiological states. Placebo responses are notably large and frequent in chronic pain treatments—i.e., placebo analgesia—presenting both opportunities and challenges. On the one hand, harnessing placebo for pain management offers opportunities for non-pharmacological, non-opioid treatments; on the other hand, strong placebo responses can obscure drug effects and complicate clinical trial design for developing analgesic drugs [1–4]. Placebo responses are thought to be driven by biological, contextual, and affective cues [5], but not everyone responds equally to placebos [6]. Thus, there is an impetus to discover the specific traits that make an individual more likely to respond to placebo. Identifying such traits and being able to predict placebo responders, could have strong implications for patient triage in clinical trials, as well as in pain management and patient care.

Previous studies have demonstrated that both neurologic factors (e.g., brain properties) and personality factors (e.g., interoceptive awareness) can predict placebo responses, but with weak generalizability and limited ecological validity [6, 7]. The latter point is crucial – since the placebo responses are thought to be driven by biopsychosocial aspects, it is unclear whether relatively stable brain properties or personality traits can capture nuances in the multiple contexts in which a patient engages with the treatment. In other words, the patient’s situational assessment of a treatment outcome is dynamic, changing in response to prior treatment experiences, coping strategies, identity, and a multitude of other psychosocial factors. Placebo responses have been shown to be impacted by these contextual contributions [5, 8], and thus novel approaches accounting for them are required. This contextual information is most readily accessible through the analysis and quantification of natural language, which can provide a wealth of knowledge about the patient’s view of self and their relationship with pain. We have previously shown that such features can be quantified using natural language processing (NLP) and that analgesic responses to placebo can be predicted with high accuracy (79%) using a model generated from NLP parameters [9, 10]. These results were later validated with an independent cohort showing that this language model could accurately predict placebo response but not treatment response, indicating generalizability and specificity to placebo [10].

Recent advancements in NLP have significantly improved the ability to capture psychosocial traits from language. Pretrained Large Language Models (LLMs) learned to encode linguistic and semantic knowledge from large amounts of diverse (i.e., linguistic styles and domains) natural language data, and can be fine-tuned to existing data to learn concepts and associations from specific domains such as pain. When analyzing a given word, their specific transformer-based architecture enforces the observation of neighboring words, embedding this context into its perceived meaning [11]. These models thus allow for extracting domain-adapted, rich semantic representations that can be leveraged for developing and interpreting a placebo response predictive model. These new methodologies overcome some limitations of past work, such as overlooking linguistic context (i.e., neighboring words) and limited interpretability. In this study, we test our ability to predict and interpret placebo responses by developing a new predictive model based on LLMs. We will assess if this pretrained, context-sensitive model can improve the predictive ability of past models developed with simpler methods, and whether LLMs can further help with the interpretation of latent semantic topics that predict placebo responders.

## Methods

This study reports a re-analysis of data from two randomized control trials (RCTs) investigating predictors for placebo in chronic low-back pain (CBP) patients [ClinicalTrials.gov registration ID: NCT02013427]. Details of these trials were published previously [6, 7], and are presented summarily here for brevity. They included two double-blinded independent studies in CBP patients, reporting pain superior to 4/10 on a visual analog scale, history of CBP for at least 6 months prior to enrollment, no pain at other body sites, and no comorbid neurological or psychological disorders. All participants stopped ongoing pain medication during the RCTs. The first study was designed to build predictive models of placebo response; and the second study was designed to validate the best model from the first study in an independent sample/study. The two studies were conducted approximately 2 years apart, and both the patients and research staff enrolled in each study were different to enhance generalizability. Both studies were approved by Northwestern University’s Institutional Review Board, and all participants signed a consent form.

These data were previously used to build predictive placebo models [9, 10] via conventional language modeling techniques including dictionary-based approaches [12] and latent semantic analyses [13], which extract semantic representations for each word independently of the surrounding text, i.e., context. The current study builds upon this previous research by developing, validating, and interpreting a model predictive of a patient’s placebo treatment response, taking advantage of contextual features extracted from their interview – that is, linguistic features which capture the meaning of a word based on the word itself and neighboring words – and using LLMs to interpret the learned model (Fig. 1). This context-sensitive feature extraction is especially important for language classification tasks where the meaning of a word can change depending on its context of use, effectively tackling challenges related with polysemy and the use of metaphors, as is frequent in the verbal expression of pain [14–16], and correctly assigning different meaning to repeated words when surrounded by different context. Additionally, words such as the pronoun “it” can be enriched with surrounding meaning, instead of being represented by a fixed embedding vector or removed as a standard stop word, disrupting the natural structure of the text [11, 17]. Thus, contextual embeddings have the potential to produce richer representations of the text, potentially leading to better predictive performance and significantly improving the interpretability of results when coupled with explainable artificial intelligence.

**Figure 1.**
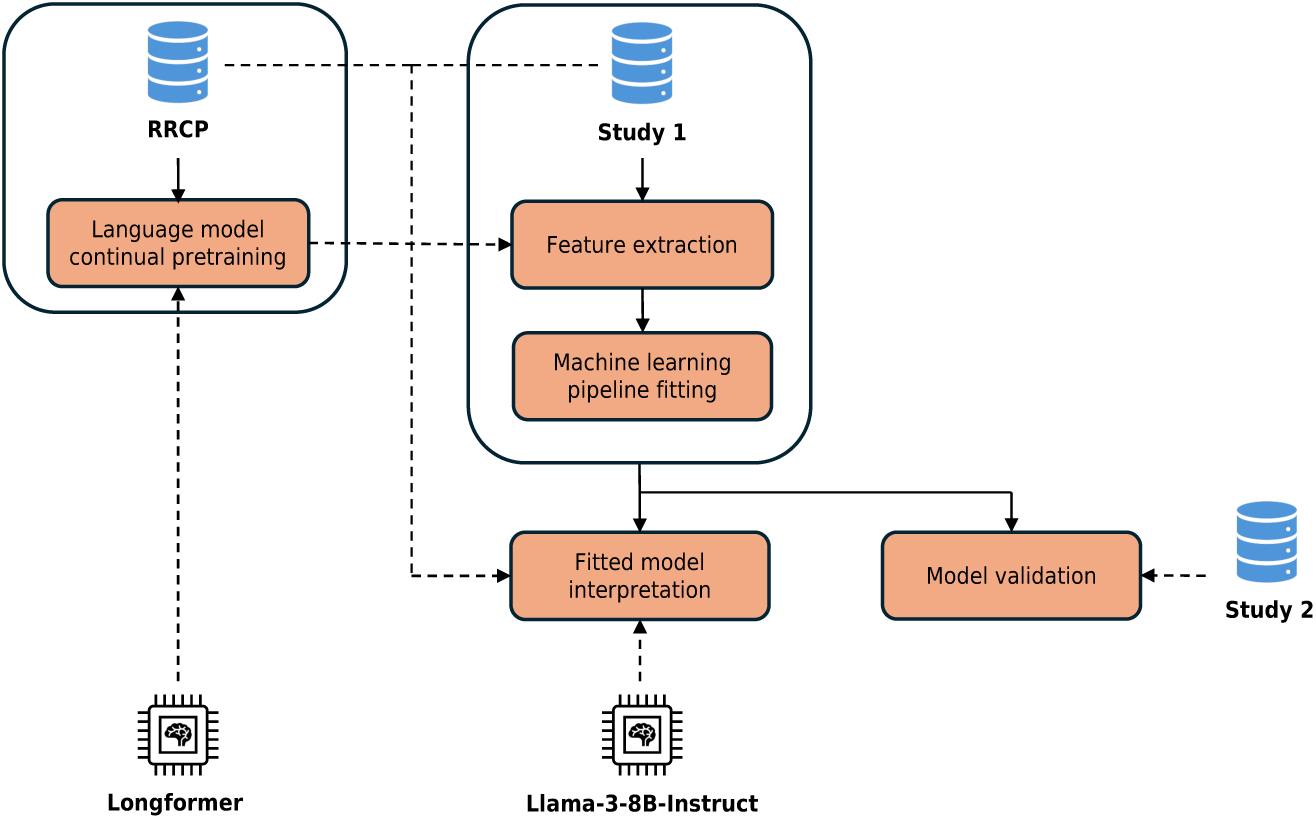
Study Design. Textual data from RRCP are used to continually pretrain the Longformer language model, for pain-domain adaptation. Textual data from study 1 are used for feature extraction, with the adapted Longformer, and fitting the machine learning pipeline, for the placebo response prediction task. This fitted model is interpreted using a variety of semantics, namely from study 1 and RRCP, leveraging the Llama-3-8B-Instruct LLM. This model is also validated on textual data from study 2.

### Participants, data, and study design

#### Study 1 participants and clinical trial design

In the first study [6, 9], 125 participants with CBP were enrolled and 66 completed all aspects of the study, including the language interviews. Full demographic data for study 1 can be found in the supplementary material. Participants were randomly assigned to one of two arms: treatment (N=46) and no-treatment (N=20). Patients in the treatment group were further split into active treatment (N=4, Naproxen, 500 mg + Esomeprazole, 20mg) and placebo treatment (N=42, 2 lactose pills) subgroups. The size imbalance between active and placebo treatment subgroups was intended for blinding purposes only, given that the goal of the study was focused on predicting placebo responders. This way, investigators could ethically tell patients they might receive a drug or a placebo without overt deception. The allocation odds were not disclosed to the participants. Since we have shown previously that the prediction of placebo responders is not confounded by regression to the mean and/or natural recovery (i.e., able to predict recovery in individuals receiving no treatment [9, 10]), in this study we focus only on predicting placebo responders within the treatment arm, and use data from the no-treatment group to help fine-tune the language model (see below).

Prior to treatment commencement, pain ratings were obtained twice daily via a smartphone app to establish baseline pain. Subjects completed 2 two-week treatment periods followed by a one-week washout. Throughout treatment and washout periods, participants continued to rate their pain twice daily. Treatment outcomes were measured during the last week of the treatment phase. To define placebo responders vs. non-responders, a permutation analysis was done for pain ratings of each subject, to assess if pain ratings during the treatment phase were lower than those from the baseline. If pain intensities for either of the treatment phases were statistically lower than baseline, patients were labelled placebo responders. Otherwise, they were labelled placebo non-responders. After the treatment and washout periods, all participants were interviewed regarding their personal experience of pain. The interview followed a semi-structured script with prompts covering 16 topics of interest (27.2 ± 10.3 minutes) but was open-ended such that the sequence of topics followed the natural flow of the conversation (see supplementary material and the original article [9] for more details).

#### Study 2 participants and clinical trial design

The second study [7, 10] enrolled 94 CBP participants, out of which 50 completed all aspects of the study. 8 participants were randomized to a no-treatment arm and 42 to the treatment arm. In this study, however, participants were *equally* randomized to an active group (N=22, Naproxen, 500 mg + Esomeprazole, 20mg) and a placebo group (N=20, 2 lactose pills). This was to investigate effect specificity and the additive nature of placebo effects over drug effects. Full demographic data for study 2 can be found in the supplementary material. As in study 1, placebo responders and non-responders were classified by permuting pain ratings from 2 weeks prior to treatment compared to the last week of treatment. However, unlike study 1, study 2 participants were interviewed about their pain experience at the start of the study, such that any predictive ability could be dissociated from treatment effects. Since study 2 was designed to replicate findings from study 1, the study’s interview script was considerably smaller than study 1 (3.27 ± 1.67 minutes), covering 4 of the original 16 topics, based on the topics that provided most predictive signal (see supplementary material and the original article[18] for more details).

#### Reddit Reports of Chronic Pain (RRCP)

Besides study 1 and study 2 data, we also used data collected from social media to fine-tune our feature extraction language model and assist with interpretability (see below). RRCP [19] is a dataset composed of Reddit posts, scraped from the web, on select subreddits (i.e., self-moderated sub-forums focusing on a specific topic). One of the included subreddits is “r/backpain,” which focuses discussion on the topic of back pain, our studied clinical population. For this subreddit, RRCP contains 2667 individual posts (from its inception up to 2020). Details about demographic, geographic, and text sentiment distributions of these posts can be found in the original paper [19]. According to Reddit’s Data API terms, academic research on the platform’s public content is permitted [20]. We did not attempt to identify or contact any Reddit user, and we do not report any identifying information. We do not publicly share any models trained on Reddit data.

### Preprocessing, data cleaning, and feature extraction

After interview transcription and quality control, the text was preprocessed, and semantic (contextual) features were extracted. For study 1, since the interview covered 16 topics in various temporal orders, each subject’s answer was automatically annotated for each of the topics, and then visually inspected and corrected for accuracy. All text was preprocessed with the same pipeline: 1) replacing all transcription annotations, e.g., “(inaudible at timestamp),” Reddit tags (subreddit and user), and URLs, with the “[UNK]” token (standing for “unknown”); 2) stripping all HTML tags, markdown styling, emojis, and multiple whitespaces; and 3) keeping all original capitalization. For study 1 only, we removed the last two topics from all interviews due to their close relation with the study itself and consequent limited generalizability (opinion of the study, and other miscellaneous topics), and the study medication topic for the same reasons, along with the fact that participants in the no-treatment arm did not answer this question. We did not remove any topic from study 2 interviews. We considered only the main body of text in all Reddit posts.

Once the text was preprocessed, segmented, and annotated, we extracted semantic representations (i.e., contextual feature vectors) for each interview from a language model, in this case, the publicly available pretrained Longformer language model [21], fine-tuned on RRCP text (i.e., domain adaptation). We chose this model for several reasons: it is open source, with proper licensing agreements; it allows the processing of larger text sequences (i.e., 4096 tokens), as opposed to other self-attention-based models, such as BERT [22] and RoBERTa [23] (i.e., 512 tokens), thus making it more adequate for long interviews (study 1: 3366.36 ± 1768.29 tokens, study 2: 450.05 ± 330.07 tokens); and it has been shown to represent long sequences of text in meaningful ways for various tasks, such as question answering and document classification [21]. We fine-tune Longformer on RRCP text, since the base model is trained on large-scale generic text and is thus domain-agnostic and possibly lacking in knowledge about pain-specific topics. We also report results using the base Longformer model for completeness.

Because interviews from both study 1 and 2 are obtained through specific topic questions, we also compared the impact of text structuring on contextual feature extraction. Thus, we structured interviews in two ways. The first, referenced as “sequential text,” assumes the original transcript sequence and represents the interview as a single document (a document is a single input sequence of text for a given patient). The second, referenced as “topic groupings” concatenates all segments of the same topic in each interview, effectively representing a single interview as a set of documents, each focusing on a single topic. Note that this approach does not change the text content; it re-arranges the text sequence based on the topic sequence such that it follows the same order across participants. In other words, while the former represents the text in line with the flow of the conversation regardless of topic coherence, the latter represents the text based on semantic information that is topic specific.

### Definition of the document feature vector

The Longformer model outputs a 768-dimensional vector for each token in the input document. We defined the document feature vector as the pooling of all token embeddings, summarizing latent semantic dimensions across the whole document. We compared two of the most common strategies to aggregate multiple vectors: mean and max pooling [16, 24, 25]. Mean pooling calculates the mathematical average of all token vectors. This approach likely dilutes sparse or extreme signals and thus reflects the overall (i.e., mean) semantic content of the whole document. This averaging potentially increases the similarity of the semantic representation vectors across subjects and therefore may generalize better, but at the cost of also averaging out predictive signal. In contrast, max pooling attributes the maximum value observed across tokens for each vector element, effectively identifying the most salient attributes of each document. This approach will identify signals in a more specific way for each interview and may be able to pick up on unique predictive signals, at the cost of being less able to identify common language patterns across subjects, lending itself to potentially weaker generalizability.

Longformer is designed for long input documents, up to 4096 tokens. However, 26% of study 1 interviews are over this limit. To avoid truncating the original text arbitrarily at the maximum length mark, for these specific cases, the tokenized document was split into two overlapping windows of 4096 tokens each, without any padding tokens, before feature extraction. Note that, in these cases, the resulting document feature vector was calculated by pooling its left and right window vectors, matching the token mean or max pooling strategy. The overlap allowed the two windows to share some of their context (i.e., neighboring words).

### Fine-tuning of Longformer

The pretrained Longformer model provides domain-agnostic contextual feature vectors. Since pain interviews employ specific morphosyntactic patterns and metaphoric references [14–16], there is reason to believe that biasing the language model towards the language of pain would result in more meaningful feature vectors (for the pain domain). This and similar techniques have been shown to improve predictive performance in similar tasks [26]. To this end, we fine-tune the Longformer model on the Masked Language Modelling (MLM) task with domain-specific text, obtained from the RRCP dataset (also known as domain adaptation). For model fine-tuning to be successful, it requires that the training data is curated to be domain-specific, such that the model learns our specific domain of pain, and a clear stopping criteria (number of training steps and parameters) to ensure the model stops fine-tuning before it starts losing pretrained knowledge, i.e., when the model starts overfitting to training data and forgets what it had learned during pretraining, also known as catastrophic forgetting [27].

To address the domain specificity of RRCP texts, since our patient sample contained only patients suffering from chronic back pain, we limited the MLM fine-tuning textual data to posts on the “r/backpain” subreddit. We further automatically filtered these posts to those discussing relevant topics of interest, according to the interview topics of study 1 (see supplementary materials for more details on the filtering algorithm). We deemed this filtering necessary to further curate the selection of RRCP texts, as we observed in-domain but out-of-topic posts, such as advertisements for websites or tools to deal with chronic back pain, and recruitment calls for back pain studies. A total of 1893 posts (out of 2667) from the “r/backpain” subreddit were used for MLM fine-tuning.

To control the amount of fine-tuning, we defined three loss functions, measured at every 50 training steps (see supplementary materials for more details). These measured how well the model predicted each masked token on three distinct validation datasets, specifically, 10% of the posts selected from “r/backpain” (190 documents), study 1 interviews from patients in the no-treatment group (20 documents; not used in the placebo treatment prediction task), and 10% of the test subset of the publicly available WikiText dataset (wikitet-2-v1; 436 documents; [28]). We expected the minimization of the loss on each of the three datasets to reflect, respectively, the desired biasing towards the pain domain, the approximation to the clinical interviews used for the predictive model development, and the retaining of the pretrained knowledge. We also used the number of frozen layers as an additional optimization parameter, balancing retaining pretrained knowledge whilst approximating the pain domain. We chose the fine-tuned checkpoint of Longformer that most minimized all three loss functions.

### Machine learning pipeline

The machine learning pipeline was selected to be identical to the best performing pipeline of our original study [9], such that current performance could be benchmarked. Specifically, it included feature scaling and selection, and a Linear Support Vector Classifier (LinearSVC) model with L1 regularization. All base models were obtained from sklearn [29]. Feature scaling and selection leveraged the RobustScaler and SelectKBest implementations, respectively. Our original study included additional variations of models and regularization [9], for which we also present results, for completeness, in the supplementary materials.

This pipeline was fitted to study 1 features through nested cross-validation (CV). The inner CV, implemented with sklearn’s GridSearchCV, performed hyperparameter optimization in 10 stratified folds, specifically searching for the best combination of the number of features to select (parameter k in SelectKBest) and the amount of model regularization (parameter C in LinearSVC). The outer CV, implemented with Leave-One-Out CV, fitted and tested (with the one left-out sample) the machine learning pipeline with the optimized hyperparameter configuration, ensuring proper independence between training and validation samples. We measured performance based on accuracy score, since the data is well balanced. We compared performance against the null distribution by permuting the labels and assessing the likelihood of the prediction accuracy occurring by chance. We tested models for four different iterations, as mentioned above: 1) mean pooling and sequential text; 2) max pooling and sequential text; 3) mean pooling and topic grouping, and 4) max pooling and topic grouping.

As in our previous study [10] the best-performing model (from the four iterations described above) was independently validated using data from study 2. To do so, the best performing model was refit in study 1 and validated on study 2. A key methodological challenge here lies in the differences between the two studies: study 2 interviews were shorter and covered only 4 of the 16 topics assessed in study 1. In our earlier work [10], a model trained on study 1 data validated successfully on study 2 data, despite differences in interview length. This was likely possible due to the methodology (i.e., averaging signals across all tokens in the interview), capturing broad themes more amenable to generalization. Here, however, the methods are more context-sensitive, and predictive features may depend on information not available in study 2 interviews. To address this, we implemented two separate validation approaches: first, we applied the model “as-is” to study 2; second, we retrained the best performing model only for the 4 overlapping questions in study 1, allowing for hyperparameters to be tuned, and validated this model with study 2. Results from both approaches are reported.

### Interpretation

One major disadvantage of deep learning feature vectors (e.g., the 768-dimensional contextual feature vectors used in our machine learning pipeline), is the difficulty in interpreting what kind of semantic or linguistic signal each dimension is capturing, if not multiple signals at once. This lack of real-world anchors hinders model interpretability and the acquisition of new knowledge about the underlying task. Input perturbation methods, such as LIME [30] and SHAP [31], target these challenges but fall short of providing novel, concrete, and actionable knowledge about the underlying task. In this work, we adopted two approaches to interpreting machine learning selected features. The first assessed associations between the extracted features and previously established placebo predictive features and linguistic variables. The second focused on further exploring the semantic space used by the fitted model, understanding the fitted decision boundary that separated placebo responders from non-responders and which semantic concepts lied on either side of that boundary. This has the potential to uncover concepts that the fitted model deemed “placebo responder”-like and “placebo-non-responder”-like but may not have otherwise been seen in correlational or qualitative thematic approaches.

#### Associations with psychological and demographic features

To begin interpreting the contextual features captured by Longformer and later selected by the machine learning pipeline, we correlated each of these features with linguistic and demographic variables, clinical questionnaires, and psycholinguistic assessments, which had been explored in previous works [6, 7, 9, 10]. We first investigated whether the features were associated with patient age, verbosity (i.e., the number of words in the interview), and vocabulary (i.e., the number of unique words in the interview), to ensure our predictions were not driven by lower-level linguistic information. Then, we assessed if the features were associated with other psychological features shown to predict placebo in our previous work. To do so, we selected relevant clinical questionnaire subfields, namely “openness” (from the big 5 personality dimensions), and 4 subscales from the Multidimensional Assessment of Interoceptive Awareness (MAIA): “Not Distracting” (MAIA/nd), “Attentive Regulation” (MAIA/a), “Emotional Awareness” (MAIA/e), and “Self-Regulation” (MAIA/sr). These were identified by Vachon-Presseau et al. (2018) as the features most correlated to the magnitude of placebo response. Finally, we studied the relationship between current features and prior semantic features; this included psycholinguistic measures like “drives”, “achievement”, and “leisure” from Linguistic Inquiry and Word Count (LIWC) [12], as well as semantic distance (SD) measurements to target topics, namely, “magnify”, “afraid”, “fear”, “awareness”, “loss”, “identity”, “stigma”, and “force”. These were identified by Berger, et al. [9] as the most significant features for placebo treatment response classification in their work.

#### Exploration of semantic concepts within the contextual feature space

The machine learning pipeline described above, once fitted to study 1 interviews, defines a hyperplane (i.e., decision boundary) separating the target classes in the contextual feature space. The farther a data point is from this boundary, the more confident the model is in its prediction (positive or negative). Because the contextual feature space encodes latent semantics in any given string of text, one possible approach to interpret that space and the decision boundary is to project a series of curated concepts (each represented by one or multiple strings of text) and measure their distance to that boundary. In other words, new strings of text can be passed through the language model feature extractor—producing semantic feature vectors— which can then be projected to our decision space and inform how a given concept is interpreted by our classification model. The farther a concept (e.g., “skepticism”) is from the decision boundary, the more confident the machine learning pipeline is in it being “placebo-responder”-like, or “placebo-non-responder”-like. Distances greater than zero classified concepts related to placebo responders, whilst distances smaller than zero classified concepts related to placebo non-responders.

We defined the set of concepts (or semantics) in three ways. First, we assessed how concepts hypothesized to be related to placebo would map onto our semantic space. To do so, we selected a set of 261 a-priori defined words, such as “wonder,” “anxiety,” “resilient,” and “hope,” that were used as predictive features in our original study [9]. Second, to explore topics used by the participants without restricting them to pre-defined words, we defined the second set of semantics according to study 1 interview topics. To this end, we extracted all individual sentences from study 1 interviews, their corresponding feature vectors, and clustered them in the feature space, using HDBSCAN [32], a hierarchical unsupervised clustering algorithm. We expected individual sentences to most likely focus on a single latent concept, and be more contextualized than individual words (e.g., “anxiety” versus “I really had no anxiety,” or “my anxiety was through the roof”). We leveraged hierarchical unsupervised clustering in two ways: (1) to group all sentences focusing on the same or similar latent concept, defining a unit of semantics, and (2) to discard all individual sentences that could not be clustered according to the algorithm, filtering the analyzed semantics only to those most discussed (curation). Thus, the second set of semantics was defined as the list of sentence clusters found in study 1. We prompted Llama-3-8B-Instruct [33], a publicly available LLM, to label each cluster of sentences with a short description (see supplementary materials for prompting details), for ease of interpretation of the latent concept represented by the corresponding cluster of sentences. LLMs have been noted for their summarization capabilities and further shown that model prompting is the key for better results in similar tasks, as opposed to model size, justifying the use of a cheaper and “smaller” 8 billion parameter model [34]. Notably, although not fixed a-priori, the scope of this second set of semantics was limited by study 1 interview topics. Third, we wanted to add chronic pain text concepts from novel, unconstrained text to further help understand the model’s boundaries. To do so, we performed the same clustering method as mentioned above but with the list of sentences extracted from “r/backpain,” from RRCP. In this case, the semantics were determined and controlled independently of this work, which we expected to provide novel insights into what semantic properties the model was using to classify placebo responders vs. non-responders.

## Results

### Study 1 and 2 samples and the placebo response

This article presents a re-analysis of data from two RCTs studying placebo responses to an inert pill in patients with CBP. In the first study, 66 patients completed all procedures, including a language interview. Of these, 4 received active treatment for blinding purposes and were therefore excluded, leaving a final sample of 62 patients with CBP (mean age = 45.3 ± 2.3 years). Among them, 20 patients were assigned to a no-treatment group, while the remaining 42 were assigned to the placebo group. In the placebo group, 23 patients (∼55%) reported significant pain relief (a meaningful change from baseline) and were classified as placebo responders, while 19 patients (∼45%) did not experience such relief and were classified as non-responders. In the second study —conducted over a year later with a different set of patients— 46 patients completed all procedures. Four patients were assigned to a no-treatment group and were excluded from the analysis. Of the remaining 42 patients (mean age = 45.3 ± 2.3 years), 20 were randomized to receive a placebo and 22 to receive naproxen + esomeprazole (active treatment, i.e., drug group). This manuscript focuses exclusively on the placebo group to avoid biases with active drug effects [10]. Of the 20 participants receiving a placebo pill, 8 (∼43%) were responders and 12 (∼57%) were non-responders.

### Placebo response prediction

This study included a comparison of the pooling strategies to obtain document contextual feature vectors from token vectors (as outputted by the fine-tuned Longformer) –mean vs. max pooling–, and the comparison of text representation strategies –sequential vs. by topic, for a total of four feature extraction configurations. For conciseness, here we describe only the results with the mean pooling strategy and sequential text representation, since this was the best performing configuration. We also present the results for the same experiment configuration using contextual features from the baseline pretrained Longformer model (i.e., not fine-tuned). The results for all other feature extraction configurations and alternative machine learning pipelines are shown in supplementary materials.

With the mean pooling strategy and sequential text representation, our model achieved statistically significant accuracies of 72% and 74%, in the inner and outer CV, respectively (outer CV: p=0.023, bootstrapped 95% CI [60, 93]). Fig. 2.A shows these results in relation to the null-hypothesis distribution, and Fig. 2.B shows the corresponding outer CV (Leave-One-Out) confusion matrix. Note that Fig. 2.A shows the same metrics for the baseline Longformer model, for comparison (non-statistically significant accuracy of 67% on the outer CV: p=0.084, bootstrapped 95% CI [62, 93]), providing further evidence for the hypothesized need for fine-tuned domain adaptation of the contextual feature extraction method.

**Figure 2.**
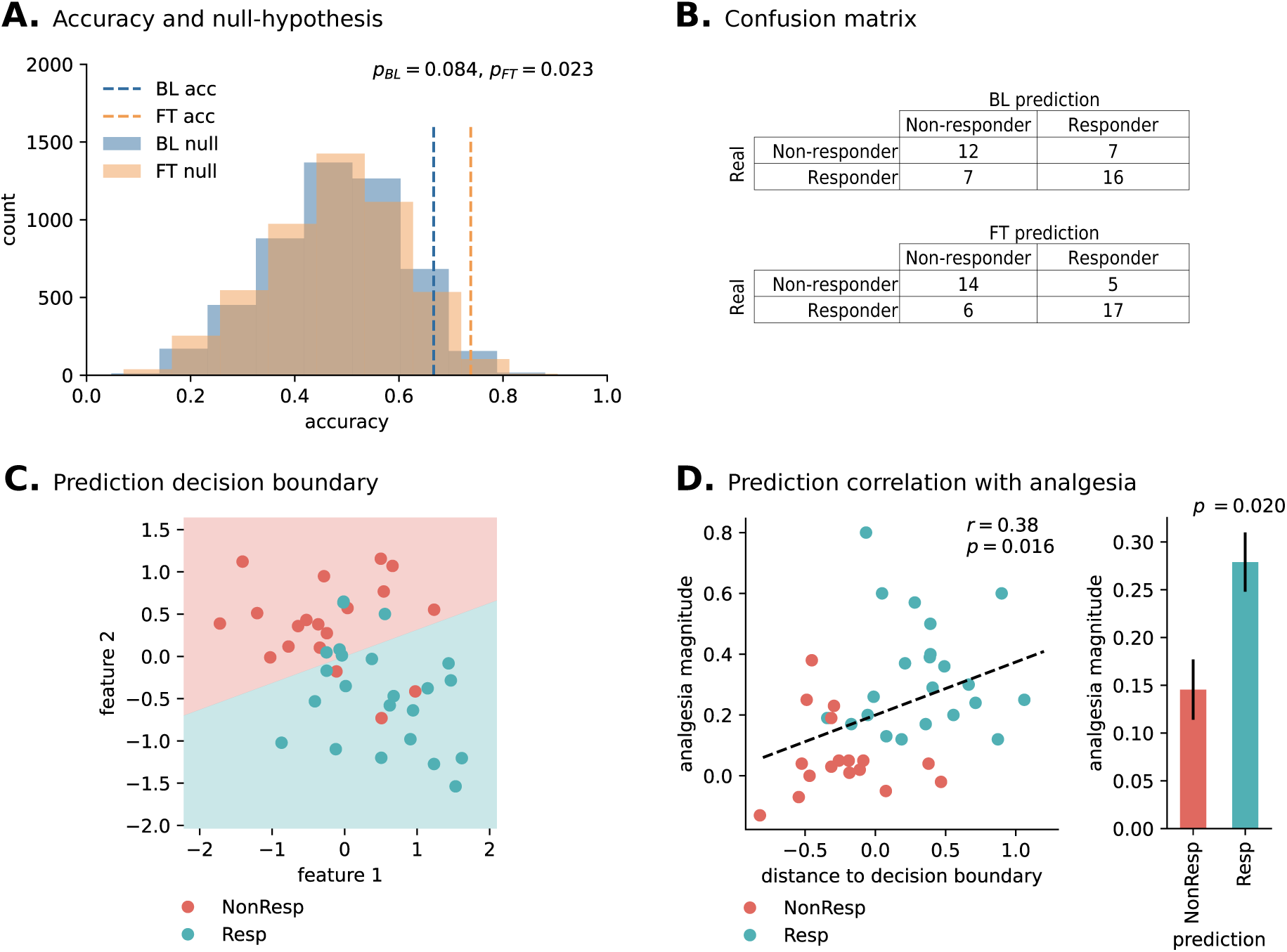
Predictive model performance. (A) Outer cross-validation accuracy based on the baseline (BL) and fine-tuned (FT) Longformer models for feature extraction, for the mean pooling and sequential text representation experiment configuration. Both results are shown in comparison to the corresponding null hypothesis distributions. The BL approach had a non-statistically significant accuracy of 67% (p=0.084), whilst the FT approach had a statistically significant accuracy of 74% (p=0.023). (B) The corresponding outer cross-validation confusion matrices. (C) Fitted decision boundary, in the selected two-feature space, in relation to the actual study 1 patient labels. Background colors represent the learned class separation, whilst the dots on top represent the actual study 1 placebo treatment subgroup patients. Red dots over red background and blue dots over blue background represent accurate predictions. (D) Relation between study 1 placebo treatment predictions and analgesia magnitude. On the left scatter plot, the analgesia magnitude is significantly positively correlated (r=0.38; p=0.016) with the distance to the decision boundary (i.e., confidence in the prediction), as shown in panel A. The right bar plot shows the expected analgesia magnitude of predicted placebo treatment outcomes. The difference between the two groups is statistically significant (p=0.020).

### Relation between prediction and analgesia

For model interpretation, we took the mean pooling and sequential text representation experiment configuration and observed the parameters of the machine learning pipeline fitted on all study 1 interviews (with hyperparameter optimization). This model had 72% accuracy in the 10-fold stratified GridSearchCV, and 79% accuracy on all training samples. Moreover, the machine learning pipeline feature selection step discarded all but two contextual features, out of the 768-dimensional feature space, which we reference as feature 1 and feature 2. Fig. 2.C shows the model’s fitted decision boundary in relation to features 1 and 2, as well as study 1 interviews and their true labels (dots). In this panel, red dots over red background (placebo non-responders) and blue dots over blue background (placebo responders) represent accurate predictions. Also, the farther a dot is from the decision boundary, the more confident the prediction. Fig. 3.D shows a statistically significant positive correlation between this distance and the actual analgesia magnitude (r=0.38; p=0.016), as well as the expected average analgesia magnitude of study 1 placebo patients predicted as responders (27,9% pain relief) and non-responders (14.6 % pain relief). The difference in analgesia magnitude between these two groups was statistically significant (independent samples t-test, p=0.020), in other words, individuals predicted as placebo responders did, indeed, show significantly higher levels of pain relief.

**Figure 3.**
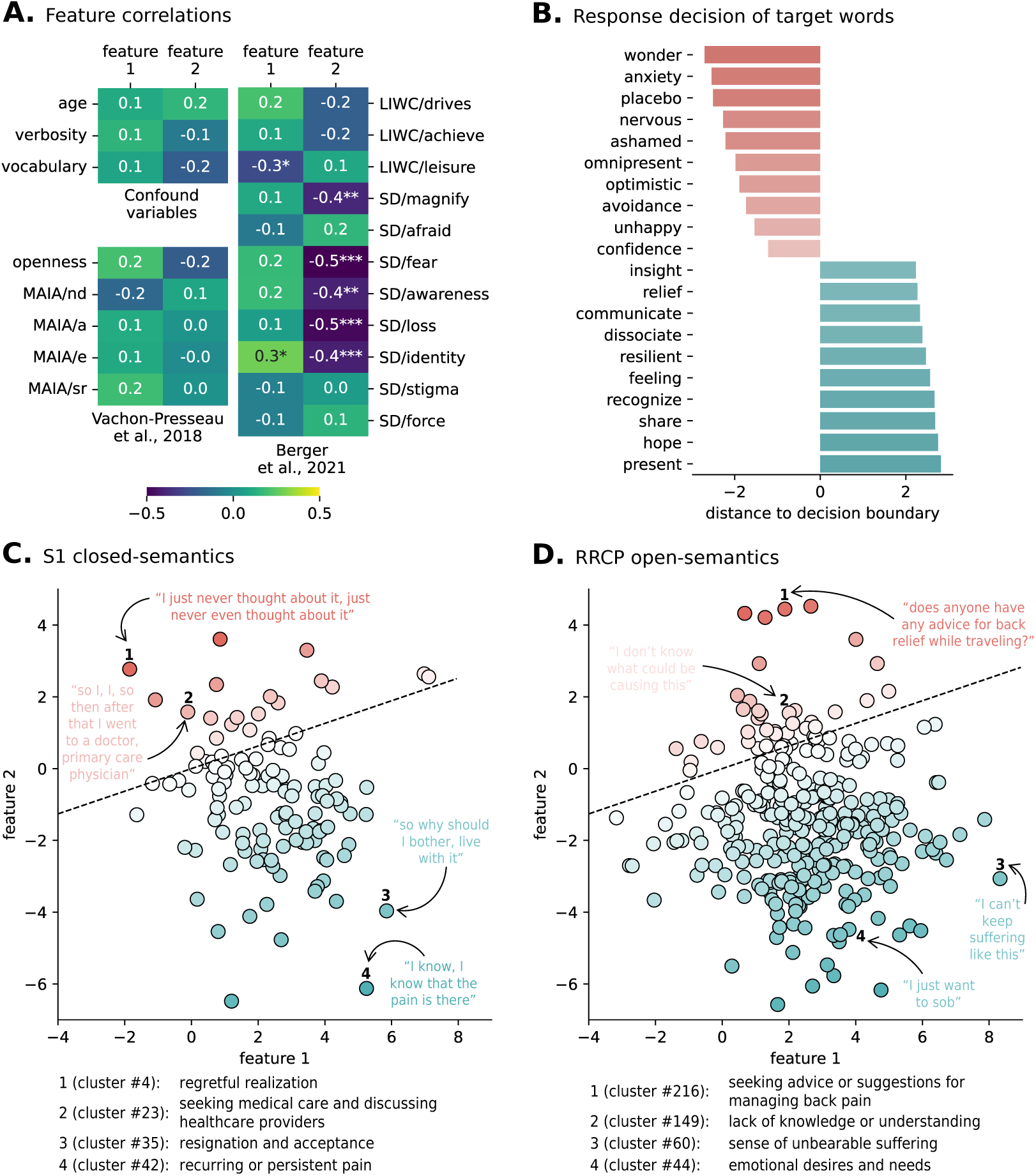
Predictive model interpretation. (A) Pearson correlation between the two selected contextual features (identified as feature 1 and 2) and other variables of interest; * p < 0.05, ** p < 0.01, *** p < 0.001. (B) Top and bottom 10 distances of words of interest to the fitted decision boundary, out of 261 words defined in previous works [9]. Red gradients represent placebo non-responder predictions, whilst blue gradients represent placebo responder predictions. (C) Scatter plot of all sentence clusters found with study 1. Dashed line represents the learned decision boundary. Red gradients represent placebo non-responder predictions, whilst blue gradients represent placebo responder predictions. Below, the automatically generated cluster description for two placebo non-responder cluster predictions and two placebo responder cluster predictions. (D) Scatter plot of all sentence clusters found with “r/backpain”. Dashed line represents the learned decision boundary. Red gradients represent placebo non-responder predictions, whilst blue gradients represent placebo responder predictions. Below, the automatically generated cluster description for two placebo non-responder cluster predictions and two placebo responder cluster predictions.

### Anchoring of contextual features on psychological and demographic features

To further understand the features selected by the classification model, and the possible redundancy with our previous results, we studied the association between the two selected contextual features (features 1 and 2, mentioned above) from our model and linguistic, demographic and clinical variables which had been explored in previous works [6, 7], as well with the language features identified in our past work [9, 10]. Fig. 3.A shows that the two contextual features do not significantly correlate with any of the confounding variables, nor the selected clinical questionnaire assessments from [6]. Feature 1 significantly correlates with LIWC/leisure (r=-0.266; p=0.037) and SD/identity (r = 0.281; p=0.027). Feature 2 significantly correlates with SD/magnify (r=-0.389; p=0.002), SD/fear (r=-0.53; p=0.000009), SD/awareness (r = −0.404; p = 0.001), SD/loss (r = −0.477; p = 0.00009), and SD/identity (r = −0.414; p= 0.0009). Thus, it seems that one of the two features (feature 2) picked up by the fine-tuned Longformer model does indeed map to similar topics as in the original manuscript. Interestingly, the other feature (feature 1) seems to capture mostly new semantic dimensions; to explore these dimensions further, we ran additional post-hoc analyses reported below.

### Semantic concept distribution over the fitted decision boundary

Fig. 3.B shows the top 10 positive and negative distances of the 261 words of interest in the first semantic set to the fitted decision boundary (shown in Fig.2.C). Like panels C and D of the same figure, the more negative the value (red gradient), the more confident the non-responder prediction, and the more positive the value (blue gradient), the more confident the responder prediction. Fig. 4.C shows the complete list of study 1 sentence clusters (each is a semantic concept represented by its sentences), plotted in reference to features 1 and 2. Each cluster is represented by a single dot, colored according to its distance to the decision boundary. The cluster position and distance to the decision boundary were calculated according to the vector averaging all included sentence feature vectors. Four clusters, two on each side of the decision boundary, were highlighted for illustrative reasons. These are accompanied by the corresponding Llama-3-8B-Instruct short description, for ease of interpretation. Fig. 3.D shows the same results for the complete list of “r/backpain” sentence clusters. Note that 124 clusters were automatically found in study 1 interviews, whilst 324 were found in “r/backpain”, which is in line with our hypothesis of unconstrained semantic concepts found in an independent sample of chronic back pain texts. These two plots show an overlap in the 2-feature space, as well as clusters in regions unique either to study 1 semantics or “r/backpain” semantics. Although the contents of only 8 clusters are illustrated here, all clusters, their corresponding clustered sentences, and short descriptions can be found and explored online at www.tinyurl.com/placeboLLM. To assess the quality of the cluster descriptions automatically generated by Llama-3-8B-Instruct, we requested 3 random raters to score their perceived level of fit of 10% of the descriptions and corresponding sentence clusters (10% of study 1 clusters and 10% of “r/backpain” clusters, randomly chosen, for a total of 44 clusters). The raters were independently asked to give their opinion on how much they agreed with the generated description, given the cluster of sentences, on a Likert scale from 0 (“I do not agree at all”) to 7 (“I completely agree”). We found an average perceived level of fit of 6.11 (CI 95% [5.83, 6.38]), with a moderate level of average inter-rater agreement, as indicated by an ICC(2,k) of 0.53 (p = 0.000547). From this, we conclude that the automatically generated cluster descriptions are, on average, perceived as a good fit for their corresponding cluster sentences. Note these labels are only used for summarization, and the full content of each cluster can be inspected in the link above.

**Figure 4.**
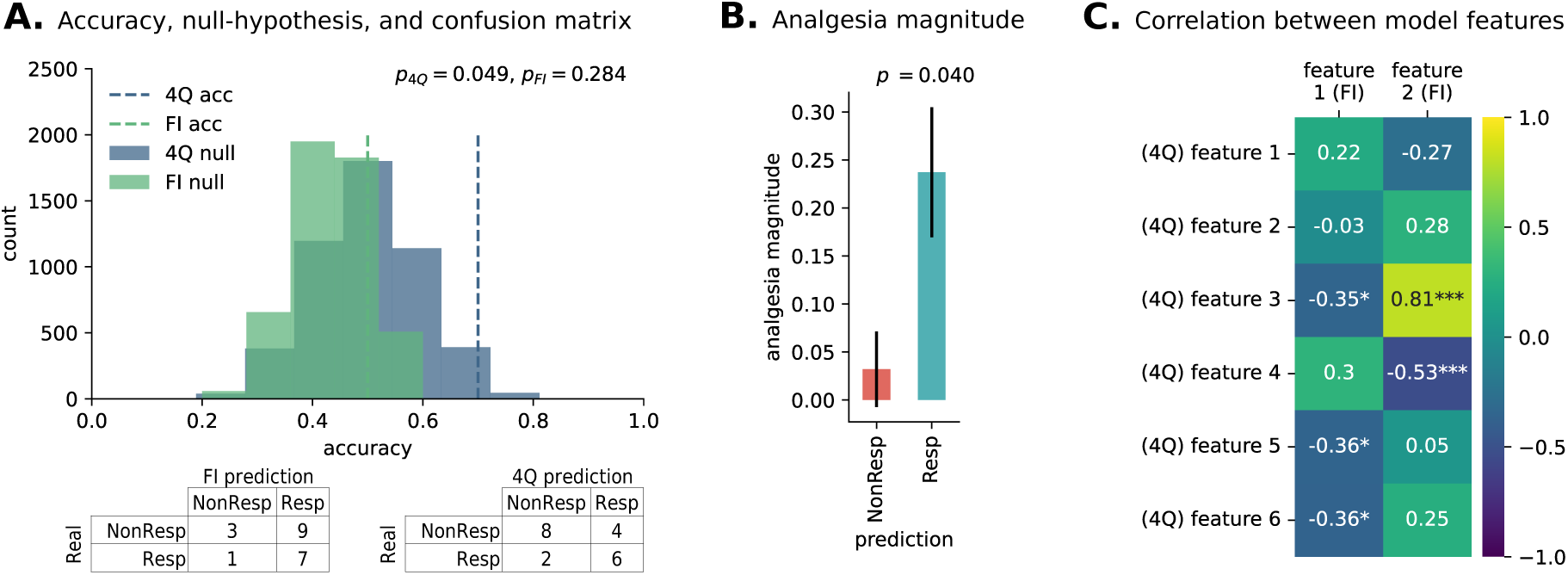
Model validation on study 2. (A) Model validation stage leveraging study 1 interviews for model fitting (single CV with hyperparameter optimization) and study 2 interviews for model testing, comparing two validation approaches. The full interview (FI; i.e., using all 16 topics in study 1 interviews) validation approach had a non-statistically significant prediction accuracy of 50% (p=0.284). The 4-question (4Q; using only the 4 matching topics from study 1 interviews) validation approach had a statistically significant prediction accuracy of 70% (p=0.049). Both results are shown in comparison to the corresponding null hypothesis distributions. Below, the corresponding study 2 confusion matrices. (B) Expected analgesia magnitude of predicted placebo treatment outcomes in study 2, based on 4Q model predictions. The difference between the two groups is statistically significant (p=0.040), highlighting that patients predicted a-priori as being placebo responders indeed show higher levels of pain relief. (C) Correlation between model features from the FI model (total of 2 significant features) and the 4Q model (total of 6 significant features) shows that while one of the FI features overlaps well with the features from the 4Q model (feature 2), the other FI feature (feature 1) is weakly correlated to features from the 4Q model. In other words, the two models capture a mix of overlapping and unique signals, likely due to differences in the interview length and content and possibly explaining why the FI model may not validate successfully in the study 2; * p < 0.05, ** p < 0.01, *** p < 0.001.

### Model validation on study 2

Thus far, the models discussed above were fitted and validated on study 1 interviews. Like our previous study [10], we also attempted a validation on an independent sample of patients, for which we used study 2 interviews. The idiosyncrasies of study 2, namely a much shorter interview script (4 out of the 16 original interview topics), and our model’s potential dependence on context prompted us to test two different validation approaches: by testing the full model (i.e. trained on the whole text) on study 2, or by instead refitting the best performing model only on the four questions that overlap between the two studies. This was decided a-priori. When testing the full interview model from study 1, the model did not validate in this out-of-sample dataset (Fig. 4.A, 50% accuracy; p=0.284) However, when the model is trained on the four overlapping interview topics only, it did validate in study 2 (Fig. 4.A, 70% accuracy; p=0.049), identifying six latent features with predictive ability. Since these two validation approaches yielded two different models, we examined, post-hoc, how the features extracted from both models compare. To do so, we generated a correlation matrix between the two original features, extracted with the full model interview (FI), and the six new features, extracted with the model limited to the 4 overlapping topics (4Q). Fig. 4.C shows the correlation matrix between features extracted with the two models. Indeed, one of the latent features from the FI model (feature 2) was strongly correlated with features from the 4Q model (strongest correlated feature, r = 0.81, p < .001) suggesting it is indeed tapping into similar latent semantic concepts; in contrast, feature 1 from FI shows only moderate associations with 4Q features (all *rs* < 0.36), hence likely capturing semantic concepts not present when the text is restricted to four questions only. These results are in line with our hypothesis that the full-interview model would learn to leverage context that would not be available in the test interviews (i.e., study 2), and thus underperform when such data is not available. Finally, the difference in analgesia magnitude between predicted placebo responders and non-responders, according to model 4Q, was statistically significant (independent samples t-test, p=0.040), i.e., individuals predicted as placebo responders did, indeed, show significantly higher levels of pain relief.

## Discussion

In this study, we tested the ability to predict placebo responders using contextual features from (large) language models, fine-tuned to chronic pain texts, and assessed the models’ ability to interpret latent topics that relate to placebo responses. To do so, we re-analyzed two clinical trials assessing the effectiveness of placebo for chronic low-back pain relief. In both trials, participants completed open-ended interviews, which we used to predict placebo treatment response. Our results can be summarized in three points. First, semantic features extracted with fine-tuned language models were able to predict placebo responders. Second, language modelling methods are able to leverage contextual information to predict placebo responses without the need to pre-select search terms, and while they are able to predict placebo responders in new datasets, their prediction ability and generalizability – at least with limited clinical data – is contingent on domain adaptation, similar document structure, and adequate pooling strategies. Third, the features extracted from language models, combined with a machine-learning pipeline, allow for the generation of a decision boundary space for placebo response prediction, thus enabling a more precise interpretation of underlying topics predictive of placebo, and showcasing how open-ended interviews can be used to generate unique insights into the psychosocial determinants of placebo response.

When analyzing interview data from the first study, the machine learning model was able to predict placebo responders with a significant classification accuracy of 74% in unseen data, as measured through nested cross-validation. That language properties can predict placebo responders corroborates our previous results [9], which used a fundamentally different language processing pipeline with important conceptual and methodological limitations. Conceptually, our previous work required selecting a series of predefined topics to predict from, which largely constrained the search of semantic topics. Arguably, this could have ignored semantic concepts not selected a-priori but that could have significant predictive value, thus justifying a more data-driven approach. Technically, our previous methods treated each word as an independent observation and were unable to disambiguate the meaning of each word given the surrounding context. Since describing pain and related concepts requires translating a subjective multi-dimensional experience into words, individuals often resort to using metaphorical language and other language tools to convey the significance of the pain experience [16] (e.g., “gnawing in my back,” “stabbing through my leg”). Consequently, models that are not able to capture and account for such nuanced semantics may be suboptimal, subject to linguistic biases, and therefore, their interpretability may be limited. In this work, we overcame such limitations by extracting latent semantics from fine-tuned language models, thus eliminating the need to select a-priori topics and making the model completely data-driven, and thus less dependent on—and biased by—the experimenter and the topic selection strategy. To do so, we leveraged domain adaptation by fine-tuning the Longformer language model to the back pain domain, providing it with enough semantic knowledge to extract features related to the chronic pain experience. Indeed, this model fine-tuning step improved feature extraction quality, increasing the accuracy of the base model from 67% to 74%, thus showing that fine-tuning significantly improves the model, and that social media data can be used to partially overcome limited access to clinically validated patient pain interviews. One can speculate that with larger and more standardized training data, these accuracy values could substantially improve classification metrics [35], and this remains to be tested in future studies. We note that the accuracy metrics reported here (74%), underperform the accuracy metrics obtained in our previous work, which obtained 79% out-of-sample accuracy. However, this more impartial approach offers important trade-offs regarding the ability to interpret the features relevant to predict placebo response, as will be discussed next. Nonetheless, it is notable that the reproducibility of our previous findings with a completely different methodology highlights the robustness of NLP to predict placebo responses, and the stability of such results regardless of the analysis pipeline, further lending strength to the concept that placebo responses are predictable biopsychosocial phenomena [8, 36].

Traditional machine learning and NLP pipelines use explicit knowledge- and domain-informed feature extraction steps (e.g., by calculating semantic distances to a-priori target words/concepts) as anchors for model interpretability [35]. Indeed, our previous studies explored this level of interpretation, identifying which latent semantic topics predict placebo responses, but as mentioned above, limiting interpretability to these topics only. Here, instead, we leverage language models–which encode latent semantics and syntax as features in a high-dimensional space [22]–allowing for a closer inspection of the semantic topics that explain the predictive ability of the model beyond the scope of pre-defined features. To understand the latent semantic topics that the model uses to predict the placebo response, we used several approaches. First, we examined the associations between the latent semantic features picked up by our model with demographic, clinical and psycholinguistic variables. We did this to anchor the latent space in previously analyzed variables. We found one of the latent semantic features identified by our model correlated strongly with language features extracted from our previously published work – more specifically, mapping into semantic dimensions of fear, awareness, and magnification. In contrast, a second significant latent semantic dimension showed weak to null correlations with previous signal, implying that the model was able to capture novel semantic domains that are predictive of placebo responses. To further understand these results and which semantic domains they map onto, we also examined the semantic distance between the full set of 261 words used in our previous work. To do this, we extracted the same two significant latent features from each of the 261 words and observed their distance and positioning with respect to the learned decision boundary separating predicted placebo responses from non-responses, effectively quantifying how each semantic concept would contribute to the placebo response. This approach identified a rich new set of semantic dimensions present throughout the subject interviews that could be used to predict placebo responses, most notably “anxiety”, “avoidance” and “wonder” as being strongly predictive of placebo non-responders, and “sharing”, “hope”, and “present” as predictive of placebo responders. Finally, to take advantage of the ability of our model to provide semantic representations at the sentence level (and with the added benefit of using context to aid in interpretability), we found data-driven clusters of similar sentences in patient interviews and social media, extracted the same significant latent semantic features, and observed each cluster’s distance and positioning with respect to the learned decision boundary. With this approach, we found that data-driven concepts such as regret (i.e., a cluster of similar sentences reliably automatically labelled as “regretful realization,” such as “I just never thought about it, just never even thought about it,” and “I never thought about that question”) and “actively seeking medical information” were used to predict placebo non-responders, whilst concepts such as “resignation” and “acceptance” most strongly predicted placebo responders. Taken together, our results fit in well with the results published in our previous work, but we also found novel concepts from both patient texts and an independent language analysis based on social media texts (e.g., lack of knowledge for non-responders, and sense of suffering for responders) that were picked up by the model and predicted the probability of an individual to respond to a placebo pill.

The interpretation pipeline developed in this study, contrary to standard approaches such as LIME and SHAP, allows for the discovery of concrete and actionable knowledge about the underlying task. For example, our model found that the concept “seeking advice or suggestions for managing backpain” is related with placebo non-response – for research purposes, this can be used as a seed for new RCTs to study the relation between proactivity in learning coping strategies and the placebo response. On the other hand, for clinical practice purposes, this concept can be used by clinicians to probe their patient’s likelihood of placebo response and leverage that new knowledge to devise treatment plans, a concept that has been advanced previously [37]. These can be extended to the more than 400 concepts discovered in this study (all concepts can be individually explored in the online tool: www.tinyurl.com/placeboLLM. Importantly, this novel interpretation pipeline is not limited to the placebo response prediction task. It can be similarly applied to any task attempting to learn relations between language and psychological states, and target outcomes.

Finally, an important note is that our model was validated and generalized on data from an independent study (study 2), but not without caveats. More specifically, only a model trained on overlapping topics across the two studies obtained classification accuracy above chance. This highlights a key limitation of state-of-the-art NLP techniques, which is the need for large amounts of data to be able to generalize to classification tasks where the training data does not exactly mimic the properties in the validation data. Given the differences in length and topic coverage across the two studies, our model did not generalize when trained on a larger topical context and tested on constrained context – our analysis suggested that it was expecting specific signals that were not present in the independent test sample. Interestingly, this was not an issue with our past results using simpler bag-of-words methods, possibly because the large averaging of semantic distances across hundreds of words also averaged out unique properties together with noise, perhaps lending itself to better generalization with the tradeoff of making each individual feature harder to interpret in isolation. With the growing availability of larger language models, and with larger samples, this effect may be mitigated and remains an obstacle to be pursued by future research.

Our study has limitations that need acknowledgment. First, given the limited available clinical data, we were not able to fully leverage the Transformer architecture behind language model design [11]. Specifically, given the limited data available, we could not fine-tune the classification token (i.e., [CLS]), which would be best-suited for prediction when fine-tuned; as a consequence, we effectively separated the data extraction and data prediction pipelines, by using the pooled token embeddings outputted by the language model as inputs to our machine learning pipeline, which may have impacted predictive ability. Further, we attempted to overcome the limited clinical data challenge by fine-tuning our model with social media data for back pain language domain adaptation, but this too entails limitations, in particular inherent biases from the data available in social media and Reddit (i.e., overall population of Reddit is skewed towards male-identified, under 35 years old from the US.). Another bias might arise from the fact that content typically presented in social media posts tap into a multitude of topics that were not included in our interview (i.e., it is essentially unconstrained), and individuals are more likely to use them for specific use cases (e.g., after a treatment, when pain gets worse, etc.) which may have affected the tone and content of such text excerpts. Finally, all semantic concepts predictive of placebo response are captured through data-driven methods and interpreted using both reverse inference and through correlation with other more well-established features. These need to be independently validated in studies specifically designed to assess these concepts, perhaps in a more quantitative and objective way including psychometrics and patient reported outcomes.

## Data Availability

All data produced in the present study are available upon reasonable request to the authors

## Acknowledgements

The authors would like to thank all members of the Apkarian lab for their feedback on the manuscript. This work was supported by National Institutes of Health grant R01AR074274 to Apkarian and by Portuguese national funds through the Foundation for Science and Technology (FCT), with reference UIDB/50021/2020. Diogo A.P. Nunes is supported by a scholarship granted by FCT, with reference 2021.06759.BD.

## References

1. Finnerup, N.B., et al., Neuropathic pain clinical trials: factors associated with decreases in estimated drug efficacy. Pain, 2018. 159(11): p. 2339–2346.

2. Kaptchuk, T.J., C.C. Hemond, and F.G. Miller, Placebos in chronic pain: evidence, theory, ethics, and use in clinical practice. Bmj, 2020. 370: p. m1668.

3. Tuttle, A.H., et al., Increasing placebo responses over time in U.S. clinical trials of neuropathic pain. Pain, 2015. 156(12): p. 2616–2626.

4. Vase, L. and K. Wartolowska, Pain, placebo, and test of treatment efficacy: a narrative review. Br J Anaesth, 2019. 123(2): p. e254–e262.

5. Atlas, L.Y., A social affective neuroscience lens on placebo analgesia. Trends Cogn Sci, 2021. 25(11): p. 992–1005.

6. Vachon-Presseau, E., et al., Brain and psychological determinants of placebo pill response in chronic pain patients. Nature Communications, 2018. 9(1): p. 3397.

7. Vachon-Presseau, E., et al., Validating a biosignature-predicting placebo pill response in chronic pain in the settings of a randomized controlled trial. PAIN, 2022. 163(5).

8. Horing, B., et al., Prediction of placebo responses: a systematic review of the literature. Frontiers in Psychology, 2014. 5.

9. Berger, S.E., et al., Quantitative language features identify placebo responders in chronic back pain. PAIN, 2021. 162(6).

10. Branco, P., et al., Predicting placebo analgesia in patients with chronic pain using natural language processing: a preliminary validation study. PAIN, 2023. 164(5).

11. Vaswani, A., et al., *Attention is all you need*, in *Proceedings of the 31st International Conference on Neural Information Processing Systems*. 2017, Curran Associates Inc.: Long Beach, California, USA. p. 6000–6010.

12. Tausczik, Y.R. and J.W. Pennebaker, The Psychological Meaning of Words: LIWC and Computerized Text Analysis Methods. Journal of Language and Social Psychology, 2009. 29(1): p. 24–54.

13. Deerwester, S., et al., Indexing by latent semantic analysis. Journal of the American Society for Information Science, 1990. 41(6): p. 391–407.

14. Halliday, M.A.K., On the grammar of pain. Functions of Language, 1998. 5(1): p. 1–32.

15. Malani, P.N., The Language of Pain: Finding Words, Compassion, and Relief. JAMA, 2010. 303(18): p. 1866–1866.

16. Lascaratou, C., The Language of Pain. 2007: John Benjamins.

17. Siino, M., I. Tinnirello, and M. La Cascia, Is text preprocessing still worth the time? A comparative survey on the influence of popular preprocessing methods on Transformers and traditional classifiers. Information Systems, 2024. 121: p. 102342.

18. Berger, S.E., et al., Quantitative language features identify placebo responders in chronic back pain. Pain, 2021. 162(6): p. 1692–1704.

19. Nunes, D.A.P., et al., Modeling Chronic Pain Experiences from Online Reports Using the Reddit Reports of Chronic Pain Dataset. Information, 2023. 14(4): p. 237.

20. Usage, R.D.P.D.A. Developer Platform & Accessing Reddit Data. 2024; Available from: https://support.reddithelp.com/hc/en-us/articles/14945211791892-Developer-Platform-Accessing-Reddit-Data.

21. Beltagy, I., M.E. Peters, and A. Cohan, Longformer: The long-document transformer. arXiv preprint arXiv:2004.05150, 2020.

22. Devlin, J., et al. *BERT: Pre-training of Deep Bidirectional Transformers for Language Understanding*. in *North American Chapter of the Association for Computational Linguistics*. 2019.

23. Liu, Y., Roberta: A robustly optimized bert pretraining approach. arXiv preprint arXiv:1907.11692, 2019.

24. Chen, Q., Z.-H. Ling, and X. Zhu, Enhancing sentence embedding with generalized pooling. arXiv preprint arXiv:1806.09828, 2018.

25. Reimers, N., Sentence-BERT: Sentence Embeddings using Siamese BERT-Networks. arXiv preprint arXiv:1908.10084, 2019.

26. Cossu, A., et al., Continual pre-training mitigates forgetting in language and vision. Neural Networks, 2024. 179: p. 106492.

27. McCloskey, M. and N.J. Cohen,Catastrophic Interference in Connectionist Networks: The Sequential Learning Problem, in Psychology of Learning and Motivation, G.H. Bower, Editor. 1989, Academic Press. p. 109-165.

28. Merity, S., et al., Pointer sentinel mixture models. arXiv preprint arXiv:1609.07843, 2016.

29. Pedregosa, F., et al., Scikit-learn: Machine Learning in Python. J. Mach. Learn. Res., 2011. 12(null): p. 2825–2830.

30. Ribeiro, M.T., S. Singh, and C. Guestrin, *“Why Should I Trust You?”: Explaining the Predictions of Any Classifier*, in *Proceedings of the 22nd ACM SIGKDD International Conference on Knowledge Discovery and Data Mining*. 2016, Association for Computing Machinery: San Francisco, California, USA. p. 1135–1144.

31. Lundberg, S., A unified approach to interpreting model predictions. arXiv preprint arXiv:1705.07874, 2017.

32. Campello, R.J.G.B., D. Moulavi, and J. Sander. *Density-Based Clustering Based on Hierarchical Density Estimates*. in *Advances in Knowledge Discovery and Data Mining*. 2013. Berlin, Heidelberg: Springer Berlin Heidelberg.

33. Dubey, A., et al., The llama 3 herd of models. arXiv preprint arXiv:2407.21783, 2024.

34. Zhang, T., et al., Benchmarking Large Language Models for News Summarization. Transactions of the Association for Computational Linguistics, 2024. 12: p. 39-57.

35. LeCun, Y., Y. Bengio, and G. Hinton, Deep learning. Nature, 2015. 521(7553): p. 436-444.

36. Branco, P., et al., 57C2.1Predictability of placebo responses and their clinical implications, in Placebo Effects Through the Lens of Translational Research, L. Colloca, et al., Editors. 2023, Oxford University Press. p. 0.

37. Fadnavis, S., et al., PainPoints: A Framework for Language-based Detection of Chronic Pain and Expert-Collaborative Text-Summarization. arXiv preprint arXiv:2209.09814, 2022.

