## Supplementary Materials for "Advancing the prediction and understanding of placebo responses in chronic back pain using large language models"

#### Study demographics

Tab. A.1 shows key demographic and clinical variable distributions of placebo treatment arm patients in both study 1 and study 2.

*Table A.1. Key demographic and clinical variable distributions of placebo treatment arm patients in both study 1 and study 2.*

| | Study 1 (N = 42) | | Study 2 (N = 20) | |
|---|---|---|---|---|
| | **Resp.** (N = 23) | **Non-Resp.** (N = 19) | **Resp.** (N = 8) | **Non-Resp.** (N = 12) |
| Age (years) | 46.9 ± 11.4 | 45.4 ± 14.7 | 56.7 ± 9.1 | 57.7 ± 11.9 |
| Sex (M/F) | 14/9 | 12/7 | 2/6 | 5/7 |
| Pain duration (weeks) | 204 ± 215 | 271 ± 469 | 306 ± 382 | 414 ± 680 |
| Pain at baseline (NRS) | 5.5 ± 2.5 | 5.8 ± 2.5 | 5.1 ± 1.1 | 5.9 ± 1.3 |

### Interview topic distribution

Study 1 interviews were obtained by transcribing patient answers to a predefined interview script covering 16 topics of interest. These were self-description, self-describing top four words, dream description, family/friends event description, pain experience/story, pain affecting self, pain influencers/coping, knowing others in pain, pain outlook, pain affecting mood/emotions, pain affecting cognition, support system/relationships, experience with medical systems, study medication – when applicable –, alternative therapies, opinion of the study, and other miscellaneous topics. The annotated segments of each interview are represented in Fig. A.1 (each block is an annotated segmented, colored by annotated topic), with the temporal topic variation shown in the *x*-axis. This figure is not intended to show each study 1 annotation in detail, but rather an overall representation of topic and interview flows. Note that most interviews started by strictly following the interview script, but throughout time the interviewer incorporated the natural flow of the conversation into the script, not abiding to a strict topical order. Some patients answered the same topic/question in multiple and/or separate instants of the interview. The extent to which each topic was discussed depended on the participant and the interview.

Study 2 interviews were obtained by transcribing patient answers to a predefined interview script covering 4 of the 16 topics of study 1. These were self-description, dream description, pain experience/story, and experience with medical systems.

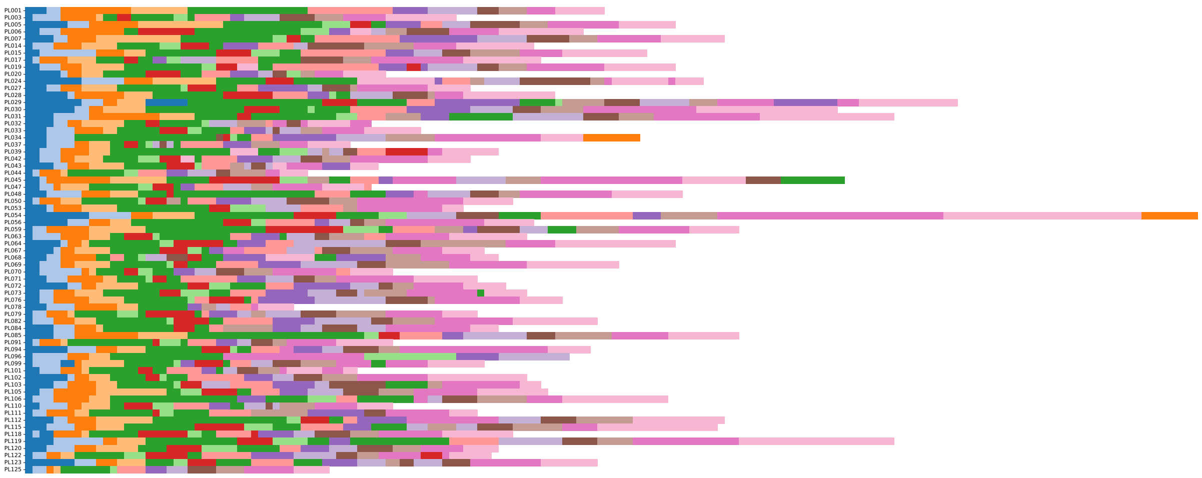

**Figure A.1.** Annotation of study 1 transcription segments, blocked by interview topic. Each pain interview is presented on the *y*-axis, and the topic variation is shown in the *x*-axis. Each patient answer is represented by a square, colored by the annotated topic. The number of patient answers (and, thus, length on the *x*-axis) does not reflect the length of the interview, as a single patient answer may have a single word or multiple words. For the same reason, the number of patient answers per topic does not reflect the length of those topics.

### Masked Language Modeling (MLM) fine-tuning

#### Textual data filtering

The Reddit Reports of Chronic Pain (RRCP) dataset contains 2667 posts in the “r/backpain” subreddit. Although the subreddit is self-moderated, there are in-domain, but out-of-scope posts advertising tools and research works that are out-of-scope for our work. Thus, we designed an algorithm to automatically filter out all out-of-scope posts (where the scope of interest was given by the topics of the study 1 interview). To this end, we concatenated all patient text pertaining to each topic (using only study 1 interviews from patients in the no-treatment arm, guaranteeing no validation data leakage into the model training routine) and used the all-MiniLM-L6-v2[1] SentenceTransformer[1] to measure the semantic similarity between each “r/backpain” post and each of the 16 topics in the study 1 interview. Semantic similarity was measured with cosine-similarity, which ranges between -1 and 1. We selected the top 2500 posts most similar to our topics of interest, according to the “elbow” heuristic (see Fig. A.2. – note that we did not choose the first “elbow” because the number of available posts would be too small for model fine-tuning, rendering it useless). We further discarded all posts with length less than the first quartile of the distribution (in this case, 78 words). In the end, 1893 posts were used for MLM fine-tuning.

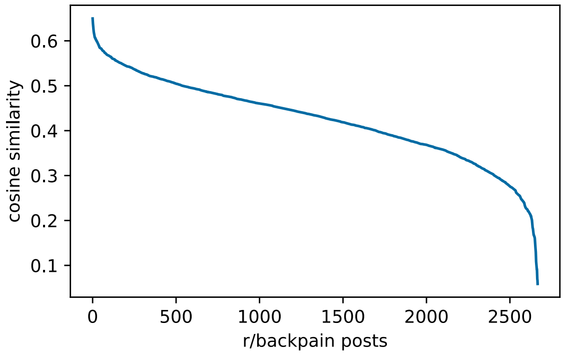


**Figure A.2.** Sorted distribution of “r/backpain” posts similarity with study 1 topics. The $y$-axis represents the cosine similarity between each “r/backpain” post and the study 1 topic with highest similarity scoring.

#### Training parameters and frozen layers

We fixed the following fine-tuning parameters according to previous related work[2] to avoid catastrophic forgetting: a learning rate of 0.00001 and 3 epochs. Moreover, we set batch size at 16, the maximum allowed by our available computing resources. At every 50 training steps (starting at zero; a training step consists of a complete backpropagation iteration over a batch of samples; note that loss scores at training step zero represent the performance of the publicly available pretrained Longformer in our evaluation setting), we measured all loss scores and saved the model checkpoint.

[1] https://huggingface.co/sentence-transformers/all-MiniLM-L6-v2

We further constrained the amount of fine-tuning by experimenting with freezing specific layers of the pretrained Longformer model. We performed a total of 7 fine-tuning experiments, all with the training parameters detailed above, where experiment *n* had the bottom *n* layers frozen (in addition to the pretrained embedding layer, which was always frozen). In deep neural networks, bottom layers are typically associated with low-level, task-transferable features, whereas top layers are associated with higher-level, abstract and task-oriented features[3]. Since Longformer is composed of 12 encoder layers, we decided to experiment freezing up to half of these layers.

#### Ranking algorithm

We measured the three loss functions, at every 50 training steps, for each of the 7 (i.e., 0 to 6 frozen encoder layers) experiments. We designed a ranking algorithm intended to find the experiment configuration which most minimized all three loss functions. To that end, given all measures, we ranked each experiment independently in each loss function, and averaged (unweighted) its ranks across all loss functions, obtaining a single ranking score per experiment.

#### Results

Fig. A.3 shows the three loss functions for the 7 experiment configurations, each referenced by the number of frozen layers, starting after the pretrained Longformer embedding layer. This figure shows that the configuration that minimizes a specific loss function is not necessarily the one that minimizes the other two loss functions, justifying the need for the ranking algorithm. The results of the ranking algorithm are shown in Tab. A.2. According to the ranking algorithm, we decided to use the MLM fine-tuned configuration with 0 frozen layers, after 100 training steps (highlighted in bold).

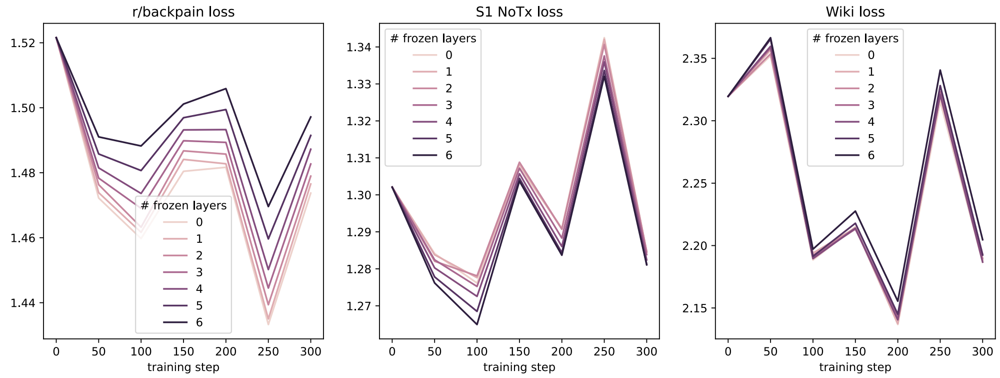


**Figure A.3.** Masked Language Modeling loss functions, by the number of frozen layers, according to the experimental setup.

*Table A.2. MLM fine-tuning ranking of experiment configurations (as identified by the number of frozen layers), at every 50 training steps.*

| # frozen layers | Training step | r/backpain loss | S1 NoTx loss | Wiki loss | r/backpain rank | S1 NoTx rank | Wiki rank | Average rank |
|---:|---:|---:|---:|---:|---:|---:|---:|---:|
| **0** | **100** | **1.459745** | **1.276148** | **2.188486** | **7** | **5** | **11** | **7.666667** |
| 3 | 100 | 1.469041 | 1.275229 | 2.190979 | 10 | 4 | 14 | 9.333333 |
| 4 | 100 | 1.473618 | 1.272548 | 2.189560 | 13 | 3 | 12 | 9.333333 |
| 1 | 100 | 1.461662 | 1.277601 | 2.194001 | 8 | 7 | 19 | 11.333333 |
| 2 | 100 | 1.463245 | 1.278094 | 2.192239 | 9 | 9 | 16 | 11.333333 |
| 5 | 100 | 1.480669 | 1.268491 | 2.191375 | 21 | 2 | 15 | 12.666667 |
| 0 | 300 | 1.473738 | 1.284626 | 2.186623 | 14 | 22 | 8 | 14.666667 |
| 1 | 300 | 1.476550 | 1.284546 | 2.188456 | 17 | 21 | 10 | 16.000000 |
| 0 | 200 | 1.481679 | 1.291008 | 2.136742 | 23 | 28 | 1 | 17.333333 |
| 6 | 100 | 1.488242 | 1.264941 | 2.197133 | 31 | 1 | 20 | 17.333333 |
| 1 | 200 | 1.482758 | 1.290694 | 2.137452 | 25 | 26 | 2 | 17.666667 |
| 4 | 300 | 1.487238 | 1.282420 | 2.186813 | 30 | 14 | 9 | 17.666667 |
| 2 | 300 | 1.478961 | 1.284668 | 2.189719 | 19 | 23 | 13 | 18.333333 |
| 2 | 200 | 1.485737 | 1.290740 | 2.140759 | 27 | 27 | 4 | 19.333333 |
| 3 | 300 | 1.482698 | 1.283853 | 2.192823 | 24 | 18 | 18 | 20.000000 |
| 3 | 200 | 1.489341 | 1.288301 | 2.143628 | 32 | 25 | 5 | 20.666667 |
| 5 | 300 | 1.491485 | 1.281241 | 2.192450 | 35 | 12 | 17 | 21.333333 |
| 4 | 200 | 1.493275 | 1.286025 | 2.140519 | 37 | 24 | 3 | 21.333333 |
| 6 | 200 | 1.505896 | 1.283678 | 2.155422 | 42 | 16 | 7 | 21.666667 |
| 5 | 200 | 1.499453 | 1.284326 | 2.144992 | 40 | 20 | 6 | 22.000000 |
| 6 | 300 | 1.497173 | 1.281085 | 2.204689 | 39 | 11 | 21 | 23.666667 |
| 2 | 50 | 1.476329 | 1.282058 | 2.353580 | 16 | 13 | 44 | 24.333333 |
| 0 | 50 | 1.472223 | 1.284124 | 2.352078 | 12 | 19 | 43 | 24.666667 |
| 1 | 50 | 1.473874 | 1.283804 | 2.356596 | 15 | 17 | 45 | 25.666667 |
| 0 | 250 | 1.433286 | 1.342499 | 2.315382 | 1 | 49 | 29 | 26.333333 |
| 3 | 50 | 1.478351 | 1.282436 | 2.358287 | 18 | 15 | 46 | 26.333333 |
| 4 | 50 | 1.481502 | 1.280262 | 2.359654 | 22 | 10 | 47 | 26.333333 |
| 5 | 50 | 1.485777 | 1.277817 | 2.365384 | 28 | 8 | 48 | 28.000000 |
| 0 | 150 | 1.480438 | 1.307450 | 2.215101 | 20 | 40 | 26 | 28.666667 |
| 1 | 250 | 1.435015 | 1.342056 | 2.320237 | 2 | 48 | 37 | 29.000000 |
| 2 | 250 | 1.439374 | 1.340647 | 2.320653 | 3 | 47 | 38 | 29.333333 |
| 4 | 250 | 1.450199 | 1.335972 | 2.320939 | 5 | 45 | 39 | 29.666667 |
| 6 | 50 | 1.491034 | 1.276185 | 2.366552 | 34 | 6 | 49 | 29.666667 |
| 3 | 250 | 1.444505 | 1.337570 | 2.323289 | 4 | 46 | 40 | 30.000000 |
| 5 | 250 | 1.459631 | 1.333637 | 2.328164 | 6 | 44 | 41 | 30.333333 |
| 1 | 150 | 1.484106 | 1.308345 | 2.214147 | 26 | 41 | 25 | 30.666667 |
| 3 | 150 | 1.489847 | 1.307209 | 2.213224 | 33 | 39 | 22 | 31.333333 |
| 2 | 150 | 1.486743 | 1.308828 | 2.213982 | 29 | 42 | 24 | 31.666667 |
| 6 | 250 | 1.469591 | 1.332078 | 2.340622 | 11 | 43 | 42 | 32.000000 |
| 4 | 150 | 1.493188 | 1.305718 | 2.213904 | 36 | 38 | 23 | 32.333333 |

| | | | | | | | | |
|---|---|---|---|---|---|---|---|---|
| 0 | 0 | 1.521565 | 1.302085 | 2.319546 | 43 | 29 | 30 | 34.000000 |
| 5 | 150 | 1.496937 | 1.304471 | 2.217881 | 38 | 37 | 27 | 34.000000 |
| 6 | 150 | 1.501145 | 1.303710 | 2.227620 | 41 | 36 | 28 | 35.000000 |
| 1 | 0 | 1.521565 | 1.302085 | 2.319546 | 44 | 30 | 31 | 35.000000 |
| 2 | 0 | 1.521565 | 1.302085 | 2.319546 | 45 | 31 | 32 | 36.000000 |
| 3 | 0 | 1.521565 | 1.302085 | 2.319546 | 46 | 32 | 33 | 37.000000 |
| 4 | 0 | 1.521565 | 1.302085 | 2.319546 | 47 | 33 | 34 | 38.000000 |
| 5 | 0 | 1.521565 | 1.302085 | 2.319546 | 48 | 34 | 35 | 39.000000 |
| 6 | 0 | 1.521565 | 1.302085 | 2.319546 | 49 | 35 | 36 | 40.000000 |

### Alternative machine learning pipelines

We studied the impact of a contextual feature extraction approach for the prediction results of the machine learning pipeline that best performed in our past work[4], i.e., feature scaling and selection, followed by the LinearSVC model with L1 regularization). In that work, three more pipelines were proposed, which vary in the classifiers and regularization, namely, with a Logistic Regression model, and L2 regularization, for which we also present here the results following our approach, for completeness. Furthermore, we also compared the differences between the mean and max pooling strategies, and the structuring of interview text, i.e., sequential text versus topic groupings. Tables A.3 and A.4 show the results for all machine learning pipelines in all experimental configurations as reported in our past work[4]. Tab. A.3 shows the results using the baseline pretrained Longformer model for feature extraction, whilst Tab. A.4 shows the results using the fine-tuned version of Longformer for feature extraction, based on RRCP text. The experimental configuration discussed in the main body of text is highlighted in bold.

*Table A.3. Results for all machine learning pipelines, in all experimental configurations, with the baseline pretrained Longformer model.*

| Classifier | Regularization | Interview structure | Pooling strategy | Inner CV accuracy (%) | Outer CV accuracy (%) |
|---|---|---|---|---|---|
| **LinearSVC** | **L1** | **Sequential text** | **Mean** | **72** | **67** |
| LinearSVC | L1 | Sequential text | Max | 74 | 57 |
| LinearSVC | L1 | Topic groupings | Mean | 65 | 57 |
| LinearSVC | L1 | Topic groupings | Max | 70 | 69 |
| LinearSVC | L2 | Sequential text | Mean | 70 | 69 |
| LinearSVC | L2 | Sequential text | Max | 74 | 52 |
| LinearSVC | L2 | Topic groupings | Mean | 64 | 52 |
| LinearSVC | L2 | Topic groupings | Max | 71 | 62 |
| Logistic Regression | L1 | Sequential text | Mean | 70 | 71 |
| Logistic Regression | L1 | Sequential text | Max | 74 | 57 |
| Logistic Regression | L1 | Topic groupings | Mean | 64 | 50 |
| Logistic Regression | L1 | Topic groupings | Max | 68 | 71 |
| Logistic Regression | L2 | Sequential text | Mean | 69 | 69 |
| Logistic Regression | L2 | Sequential text | Max | 72 | 57 |

| | | | | | |
|---|---|---|---|---|---|
| Logistic Regression | L2 | Topic groupings | Mean | 63 | 40 |
| Logistic Regression | L2 | Topic groupings | Max | 70 | 62 |

*Table A.4. Results for all machine learning pipelines, in all experimental configurations, with the fine-tuned Longformer model.*

| Classifier | Regularization | Interview structure | Pooling strategy | Inner CV accuracy (%) | Outer CV accuracy (%) |
|---|---|---|---|---|---|
| **LinearSVC** | **L1** | **Sequential text** | **Mean** | **72** | **74** |
| LinearSVC | L1 | Sequential text | Max | 65 | 55 |
| LinearSVC | L1 | Topic groupings | Mean | 65 | 57 |
| LinearSVC | L1 | Topic groupings | Max | 56 | 50 |
| LinearSVC | L2 | Sequential text | Mean | 69 | 71 |
| LinearSVC | L2 | Sequential text | Max | 66 | 55 |
| LinearSVC | L2 | Topic groupings | Mean | 65 | 57 |
| LinearSVC | L2 | Topic groupings | Max | 57 | 48 |
| Logistic Regression | L1 | Sequential text | Mean | 69 | 74 |
| Logistic Regression | L1 | Sequential text | Max | 64 | 60 |
| Logistic Regression | L1 | Topic groupings | Mean | 62 | 55 |
| Logistic Regression | L1 | Topic groupings | Max | 57 | 38 |
| Logistic Regression | L2 | Sequential text | Mean | 70 | 74 |
| Logistic Regression | L2 | Sequential text | Max | 65 | 64 |
| Logistic Regression | L2 | Topic groupings | Mean | 63 | 57 |
| Logistic Regression | L2 | Topic groupings | Max | 57 | 40 |

### Short description generation with Llama-3-8B-Instruct

#### Prompt

Given a cluster of sentences, we prompted Llama-3-8B-Instruct to generate a short description of the underlying, shared topic. We used this description to reference the cluster, as a unit of semantics. The prompt, composed of a system and user instruction, is shown in Tab. A.5. We processed Llama's answers to extract only the relevant description, e.g., from "the underlying concept is rejection or refusal.", we extracted only "rejection or refusal" (without punctuation).

*Table A.5. Llama-3-8B-Instruct prompt to generate a short description of a cluster of sentences.*

| Role | Content |
|---|---|
| system | Your job is to figure out the underlying concept of a group of sentences. Try to be specific, for example, if a group of sentences talks about durations of time, specify if long or short durations. Your answer contains only a short description of the underlying concept. You always use few words. |
| user | What is the underlying concept to the following sentences? (*list of sentences)* |

#### Validation of automatic short descriptions

To assess the quality of the cluster descriptions automatically generated by Llama-3-8B-Instruct, we requested 3 random raters to score their perceived level of fit of 10% of the descriptions and corresponding sentence clusters (10% of study 1 clusters and 10% of "r/backpain" clusters, randomly chosen, for a total of 44 clusters). The raters were independently asked to give their opinion on how much they agreed with the generated description, given the cluster of sentences, on a Likert scale from 0 ("I do not agree at all") to 7 ("I completely agree"). Fig.A.4 shows the rating distribution across raters as well as the average rating between all three. We found an average perceived level of fit of 6.11 (CI 95% [5.83, 6.38]), with a moderate level of average inter-rater agreement, as indicated by an ICC(2,k) of 0.53 (p = 0.000547). From this, we conclude that the automatically generated cluster descriptions are, on average, perceived as a good fit for their corresponding cluster sentences. Fig. A.4 shows the distribution of rater scores, along with the mean score across all raters.

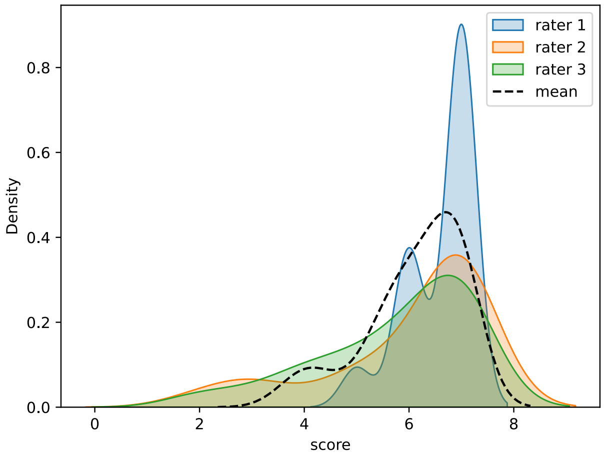


**Figure A.4.** Distribution of rater scores, along with the mean score across all raters.

### References

1. Reimers N. Sentence-BERT: Sentence Embeddings using Siamese BERT-Networks. arXiv preprint arXiv:190810084. 2019.
2. Fadnavis S, Dhurandhar A, Norel R, Reinen JM, Agurto C, Secchettin E, Schweiger V, Perini G, Cecchi G. PainPoints: A Framework for Language-based Detection of Chronic Pain and Expert-Collaborative Text-Summarization. arXiv preprint arXiv:220909814. 2022.
3. Yosinski J, Clune J, Bengio Y, Lipson H. How transferable are features in deep neural networks? Advances in neural information processing systems. 2014;27.
4. Berger SE, Branco P, Vachon-Presseau E, Abdullah TB, Cecchi G, Apkarian AV. Quantitative language features identify placebo responders in chronic back pain. PAIN. 2021;162(6).
